# Predicting COVID-19 cases across a large university campus using complementary built environment and wastewater surveillance approaches

**DOI:** 10.1101/2024.01.24.23300025

**Authors:** Aaron Hinz, Jason A. Moggridge, Hanna Ke, Alexandra M. A. Hicks, Evgueni Doukhanine, Michael Fralick, Laura Hug, Derek MacFadden, Hebah Mejbel, Caroline Nott, Ashley Raudanskis, Nisha Thampi, Alex Wong, Rees Kassen

## Abstract

**Background:** Environmental surveillance of SARS-CoV-2 via wastewater has become an invaluable tool for population-level surveillance. Built environment sampling may provide complementary spatially-refined detection for viral surveillance in congregate settings such as universities.

**Methods:** We conducted a prospective environmental surveillance study at the University of Ottawa between September 2021 and April 2022. Floor surface samples were collected twice weekly from six university buildings. Samples were analyzed for the presence of SARS-CoV-2 using RT-qPCR. A Poisson regression was used to model the campus-wide COVID-19 cases predicted from the fraction of floor swabs positive for SARS-CoV-2 RNA, building CO_2_ levels, Wi-Fi usage, and SARS-CoV-2 RNA levels in regional wastewater. We used a mixed-effects Poisson regression analysis to model building-level cases using viral copies detected in floor samples as a predictor. A random intercepts logistic regression model tested whether floor samples collected in high-traffic areas were more likely to have SARS-CoV-2 present than low-traffic areas.

**Results:** Over the 32-week study period, we collected 554 floor swabs at six university buildings. Overall, 13% of swabs were PCR-positive for SARS-CoV-2, with positivity ranging between 4.8% and 32.7% among university buildings. Both floor swab positivity (Spearman r = 0.74, 95% CI: 0.53-0.87) and regional wastewater signal (Spearman r = 0.50, 95% CI: 0.18-0.73) were positively correlated with on-campus COVID-19 cases. In addition, built environment detection was a predictor of cases linked to individual university buildings (IR_log10(copies) + 1_ = 17, 95% CI: 7-44). There was no significant difference in detection between floors sampled in high-traffic versus low-traffic areas (OR = 1.3, 95% CI: 0.8-2.1).

**Conclusions:** Detection of SARS-CoV-2 RNA on floors and viral RNA levels found in wastewater were strongly associated with the incidence of COVID-19 cases on a university campus. These data suggest a potential role for institutional built environment sampling, used together with wastewater surveillance, for predicting COVID-19 cases at both campus-wide and building level scales.

## INTRODUCTION

The coronavirus disease 2019 (COVID-19) pandemic was caused by the emergence of the severe acute respiratory syndrome coronavirus 2 (SARS-CoV-2), a highly transmissible and pathogenic coronavirus^1^. Population-scale diagnostic testing and contact tracing have been crucial tools to inform public health responses^2^. In addition, environmental surveillance strategies based on shedding of the virus in wastewater and on built environment surfaces provide information on disease burden at community and building-level scales^3–5^. These non-invasive monitoring approaches can fill the information gap left by the decline of mass individual testing and provide continual monitoring of community-wide prevalence of infections.

Wastewater-based epidemiology has successfully been used to detect the spread of SARS-CoV-2 by quantifying viral RNA levels in wastewater over time^6,3,7,8^. Wastewater-focused surveillance programs have previously been implemented on university buildings and dormitories, and have been shown to be a successful proactive tool for outbreak monitoring on university campuses^9,10^. However, correlating virus levels in wastewater with epidemiologically identified cases may be challenging due to inconsistent spatial variability and resolution^11^. Although wastewater sampling can be used to monitor individual buildings, demonstrated by its use on university campuses, there is still a gap between community- and individual-level surveillance methods.

Built environment sampling could complement these approaches by providing evidence on the presence of infected individuals at more spatially resolved scales (i.e., buildings or locations within buildings). SARS-CoV-2 transmission occurs through respiratory droplets and aerosols expelled by infected individuals, and viral particles and RNA exhibit a prolonged presence on built environment surfaces^1,12^. Detection of SARS-CoV-2 RNA from surface samples of the built environment, particularly from floor swabs^13,14^, has shown potential as a spatially resolved COVID-19 surveillance strategy^15,16^. In hospitals, significant correlations have been observed between viral loads from clinical samples and positive detection of environmental samples taken from patient rooms^17,18^. Furthermore, we have demonstrated that detection of SARS-CoV-2 on floors in long-term care facilities is strongly associated with, and may even anticipate, COVID-19 outbreaks, suggesting that floor-based sampling can play a valuable role in improving early outbreak identification^4^.

As a novel tool, the integrated use of wastewater detection with surface surveillance of the built environment may thus provide an opportunity for improved SARS-CoV-2 infection prevention and control, bridging the gap between community-scale and individual-level surveillance methods. To further evaluate this idea, we performed surface sampling at six buildings at the University of Ottawa and compared the findings to publicly available wastewater data. Our objective was to quantify the association between ambient SARS-CoV-2 in the built environment, regional wastewater signal, and near real-time COVID-19 case identifications on a university campus, a setting in which surface sampling has not been fully explored.

## METHODS

### Built environment sampling

Built environment sampling was performed in six buildings on the main campus of the University of Ottawa (uOttawa), selected with input from the uOttawa COVID-19 Recovery Task Force, a multi-stakeholder committee composed of representatives from across the university charged with maintaining health and safety of the university community during the COVID-19 pandemic. (https://www.uottawa.ca/about-us/sites/g/files/bhrskd336/files/2022-05/COVID%2019%20Recovery%20Taskforce%20TOR%20v.7.pdf). Floor surfaces were swabbed due to the reliable recovery of SARS-CoV-2 RNA compared to other built environment surfaces^4,16^. Buildings and sites were chosen to provide representative sampling of areas visited by student and staff populations and include a student residence hall (RES), the main library (LIB), and buildings hosting sports facilities (GYM), the school of management (FAC), central administration and infoservices (ADM1), and campus protection and postal services (ADM2). Two sites within each building were sampled: a high traffic site located at a main entrance or corridor, and a low traffic site at a more remote location (i.e., typically on a different level). The designation of high and low traffic site was made on the advice of building and facilities management prior to the start of sample collection. Individual classrooms were not sampled, but hallway floors outside of classrooms were swabbed in several buildings (GYM, FAC, ADM1).

Floor samples were collected twice weekly (Tuesdays and Thursdays) from September 21, 2021 to April 5, 2022. Sampling was paused after December 9, 2021, and resumed for the winter term on January 11, 2022 when the campus re-opened. To reduce the spread of the Omicron variant of COVID-19, the return to campus for the winter term was postponed from January 4 to January 31, 2022. As a result, five sampling sites at ADM1, FAC, and LIB were inaccessible until February 1, 2022.

### SARS-CoV-2 detection from floor swabs

Floor swabs were collected as previously described using a P-208 Environmental Surface Collection Prototype kit from DNA Genotek^4,16^. An approximate 2-inch by 2-inch area of floor was swabbed for 30 seconds, and swabs were immersed in nucleic acid stabilization solution and stored at room temperature until processing. Nucleic acid extractions were performed using the MagMAX Viral/ Pathogen II (MVP II) Nucleic Acid Isolation Kit (Thermo Fisher Scientific, Waltham, MA), and SARS-CoV-2 RNA was detected by RT-qPCR amplification of the N1 gene as described^4,16^. Viral copies were estimated from quantification cycle (Cq) values using the virus standard curve reported in our previous study^16^.

### University CO_2_ and Wi-Fi measurements and regional wastewater data

Carbon dioxide and Wi-Fi usage data were collected during the sampling period as proxy measures of ventilation and building occupancy. Carbon dioxide measurements were taken concurrently with each floor swab sample using a handheld digital CO_2_ sensor (Gain Express Model AZ 7755). Campus Wi-Fi activity was collected by the university as daily peak (number of simultaneously-connected) wireless devices per building. Wi-Fi data was collected for 5 of the 6 buildings sampled, excluding the residence hall (RES). Regional waste-water data were obtained from The Public Health Environmental Surveillance Database github repository (https://github.com/Big-Life-Lab/PHESD/).

### Reported cases on campus

The university administration recorded 217 self-reported COVID-19 cases between July 2020 and early February 2022. These records include one or more dates (with data missing for a subset of cases), describing the onset of symptoms (53% missing), the test date (77% missing), the positive test result date (13.5% missing), or the date when the isolation period ended (3% missing). We used the test result dates as index dates for cases. For cases where test result dates were unavailable, we filled in values using the isolation-end date minus five days, or if unavailable, then we used the symptoms onset date plus three days. These corrections were based on the mean differences between the various events where multiple events were recorded and reflect reasonable estimates of the onset of infection given data on the reported events^2^.

### Statistical analysis

Statistical analysis was performed using R version 4.2.2 (2022-10-31)^19^. Graphics were created with ggplot2 (v3.4.1). Code and data are available from our GitHub repository: https://github.com/CUBE-Ontario/UOttawa-Analysis (*to be made public upon publication*).

To examine the relationship between the university case burden and environmental surveillance variables, we fit Poisson regression models with the number of cases as the outcome. We employed backward elimination to select predictors from the aggregated results (means) of surface swabbing, waste-water testing, CO_2_, and Wi-Fi user counts. Predictors were centered and scaled to facilitate comparison of regression coefficients. Models were fit using ‘glm’ in R. The assumption of equidispersion was tested and multicollinearity was evaluated through generalized variance inflation factors.

To investigate the relationship between case incidence and surface swab results on a per-building basis, we used mixed-effects models with a random intercept for each building. First, we evaluated a logistic regression model with the presence of SARS-CoV-2 infected individuals as a binary outcome (i.e., positive event: one or more cases occurring during a week), with surface swab PCR-positivity as a fixed-effect, and with a random intercept for each building. We fit a second logistic regression model to test whether high-traffic locations contained greater quantities of viral RNA than low-traffic locations. Mixed-effects models were created using the ‘glmer’ function from the ‘lme4’ package^20^.

To model case counts at the building level, we fit a Poisson regression with a random intercept for each site, using the quantity of SARS-CoV-2 RNA recovered by PCR as a predictor. For this model, our predictor was the log-transformed mean number of viral copies (plus one) per-building during weekly intervals. Our model formula was: *cases ∼ log_10_(copies + 1) + (1|site)*. This random effects model was created using the ‘glmer’ function from the ’lme4’ package. From the fitted model, we computed incidence ratios and Wald-type 95% Confidence Intervals (CIs).

## RESULTS

### Environmental detection of SARS-CoV-2 correlates with COVID-19 cases on campus

There were 116 reported cases of COVID-19 among students and staff on campus during the study period, with case reporting ending in February 2022. Low numbers of cases were reported in the fall term, and the highest numbers reported in January, coinciding with a campus-wide closure in response to the Omicron variant (Fig. 1). The aggregate built environment surveillance trends broadly paralleled case incidence, with low SARS-CoV-2 detection during the fall term (September to December) and a spike in floor swab positivity in January (Fig. 1). A decline after the January peak preceded a second increase in environmental SARS-CoV-2 signal observed in late March (Fig. 1). These university case and built environment detection trends correspond with two increases in COVID-19 prevalence reported in Ottawa city-wide wastewater data (Fig. 1).

**Figure 1.**
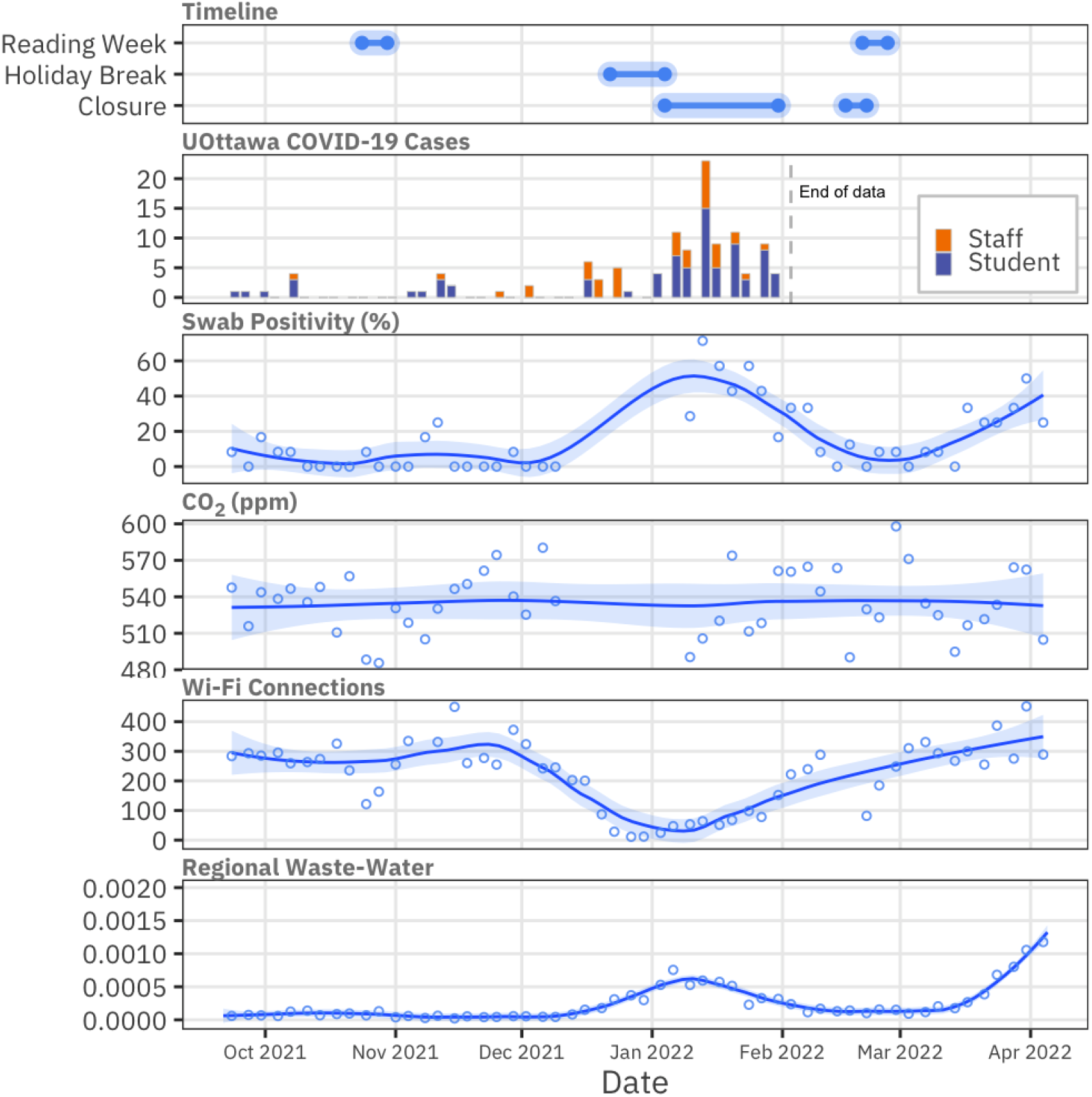
Time-series of (top to bottom): important events at the university, university COVID-19 cases, proportion of PCR-positive floor swabs, biweekly mean ambient CO_2_ (across collection sites), biweekly mean peak number of Wi-Fi connections (across 5 buildings), biweekly mean regional waste-water detection (relative to PPMoV). Points show biweekly means. Trend lines fit with the Loess method.

We observed positive correlations between university cases, swab positivity, and City of Ottawa wastewater signal (Fig. 2). The strongest correlations were between cases and swab positivity (Spearman r = 0.74, 95% CI: 0.53-0.87) and Ottawa wastewater and swab positivity (Spearman r = 0.68, 95% CI: 0.5-0.81). Neither of the building occupancy proxies (i.e., CO_2_ levels or Wi-Fi usage) correlated with campus COVID-19 cases or floor swab detection during the study period (Fig. 2). CO_2_ concentrations were stable across buildings and time (Fig. 1), at levels indicating high indoor air quality (median: 531 ppm, range: 418 - 905 ppm). Wi-fi usage, on the other hand, fluctuated during the term, with expected drops during reading week, winter break, and individual building closures (Figs. 1, 3). These results suggest that built environment surveillance could serve as a predictor for estimating active campus cases and that environmental detection of SARS-CoV-2 at the university paralleled city-wide trends.

**Figure 2.**
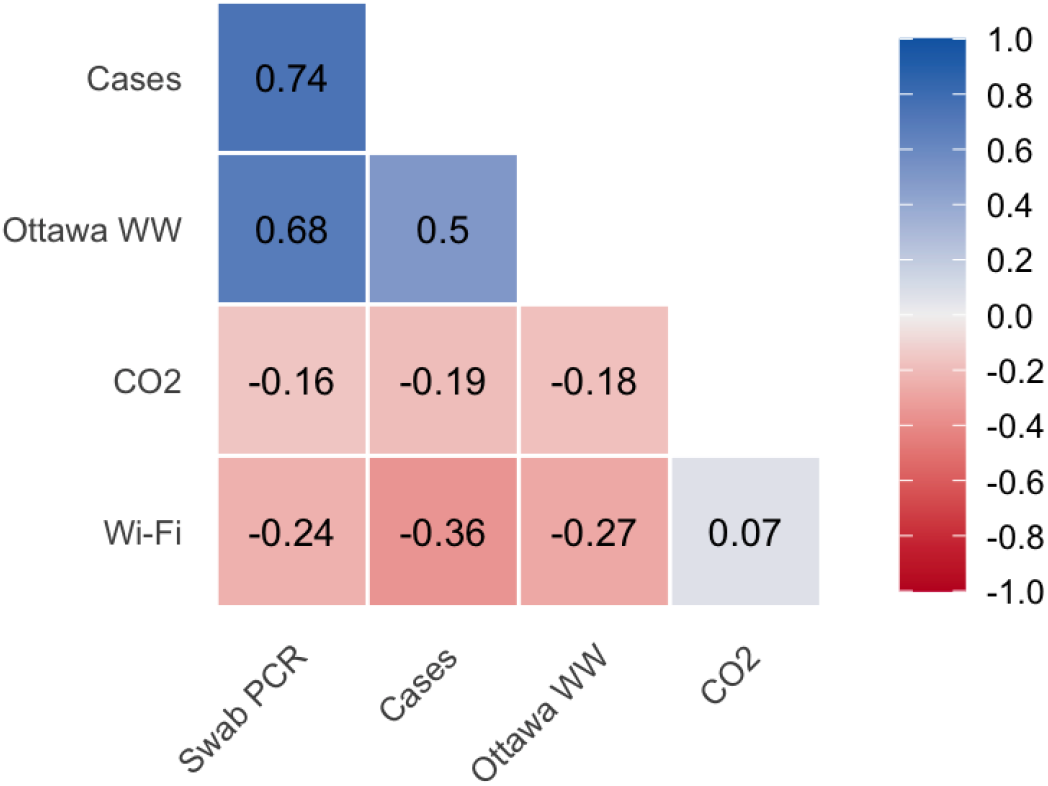
Spearman correlation between biweekly campus-wide variables: self-reported cases, floor swab positivity (Swab PCR), mean CO_2_, mean daily peak Wi-Fi connections, waste-water signal at the regional level (Ottawa WW),

### Predictive modeling of campus-wide cases based on environmental detection and building occupancy estimates

Non-invasive approaches to monitoring SARS-CoV-2 levels such as environmental sampling are valuable when systematic testing of individuals is not feasible. We evaluated predictors of the campus-wide case burden using Poisson regression models selected by backward elimination. Predictors specified in the full model included surface swab positivity (across 6 buildings), CO_2_ concentrations (at the time of swab collection), regional wastewater metrics, and Wi-Fi user counts. Backward elimination dropped the CO_2_ and Wi-Fi terms and indicated swab positivity and regional waste-water signals as significant, positive predictors of case counts (Table 1). The regional wastewater signal and swab positivity terms had similar incidence rate ratios in the final model (Table 1). The selected model did not violate the assumption of equidispersion (dispersion = 1.35, p > 0.05).

**Table 1.**
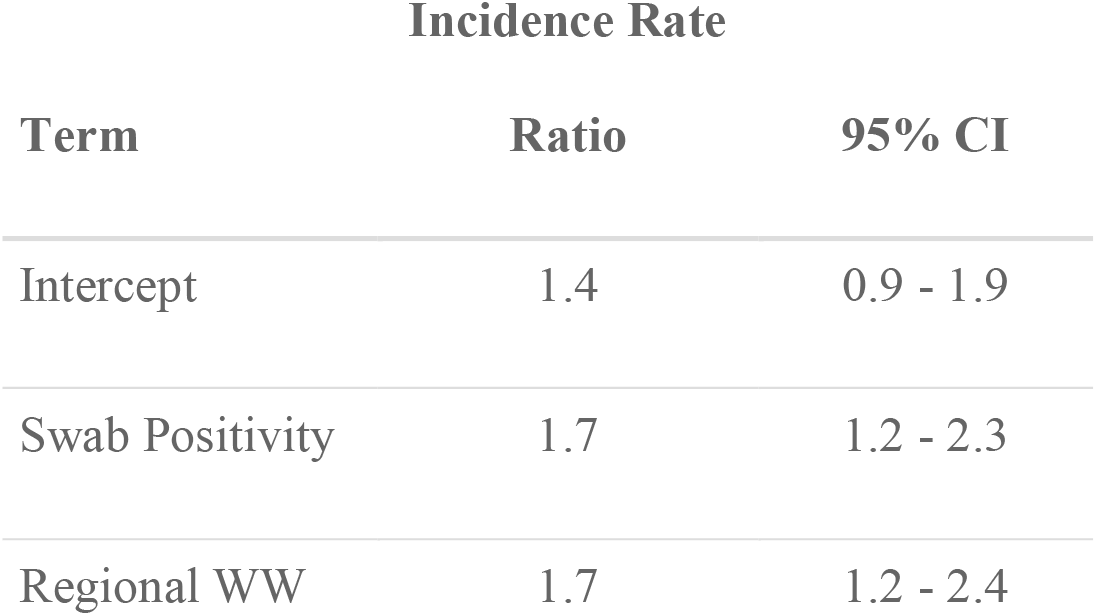
Summary of the Poisson regression model selected by backward elimination, considering main effects only (n=29).

### Modeling cases in individual buildings from built environment detection

Campus buildings visited by individuals infected with COVID-19 were self-reported during collection of campus-wide case data. We, therefore, leveraged the spatial resolution of our built environment sampling to determine whether cases in individual buildings were associated with higher levels of SARS-CoV-2 detection from floor swabs collected from the same buildings. Indeed, two sites with high positivity in January 2022 at the university residence (RES) and an administrative building (ADM2), were associated with clusters of reported cases in students and staff, respectively (Fig. 3).

**Figure 3.**
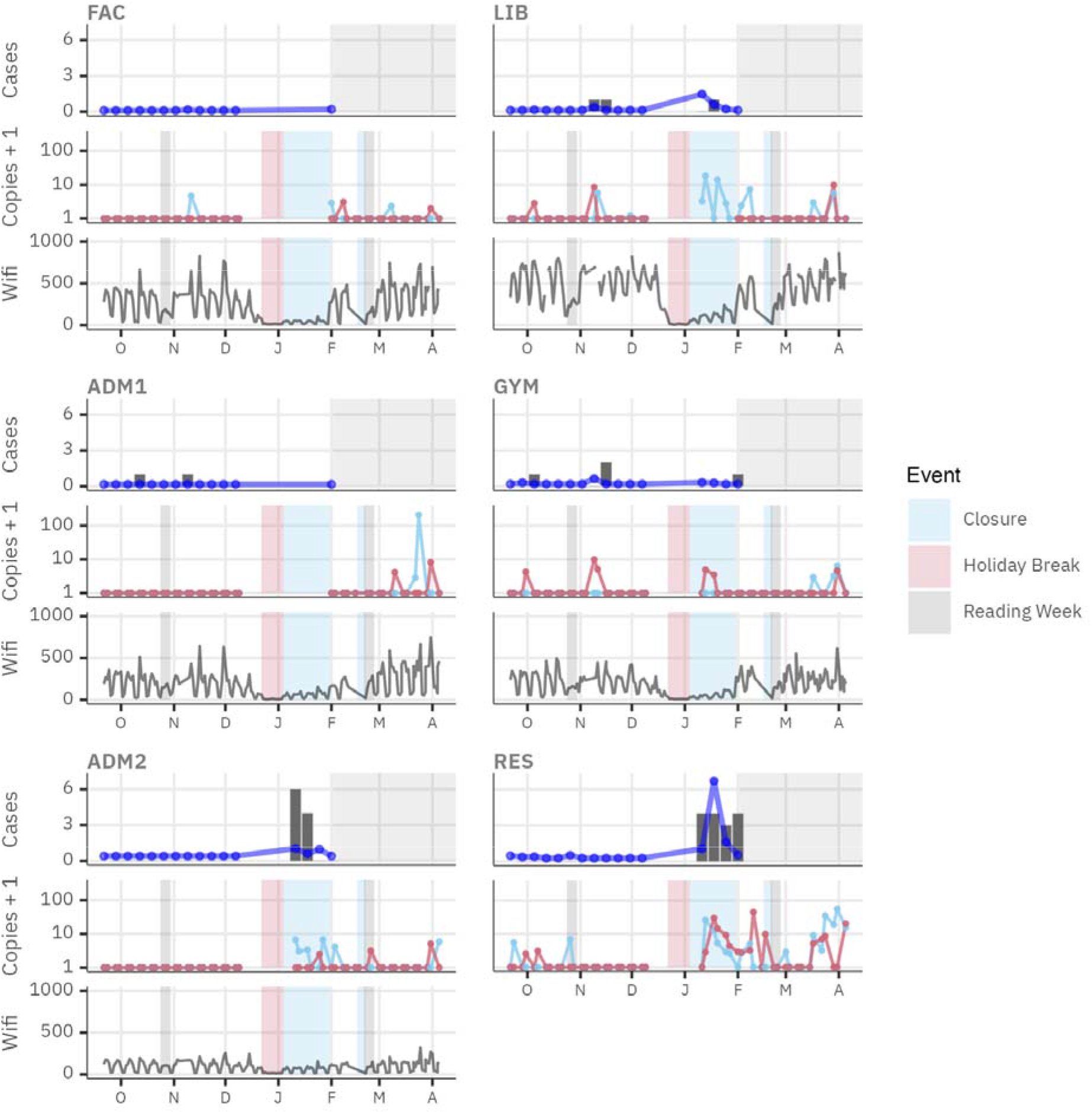
Campus COVID-19 cases, swab results, and daily peak Wi-Fi connections by building. Cases plots show counts of self-reported cases as bars and fitted values from the poisson mixed model as blue points. Case data collection was abandoned in early February 2022 (shaded area). Swab plots show results at two locations within each building, with one sample collected in high-traffic area (blue) and the other in a low-traffic area (red). Swab PCR results are expressed as the number of SARS-CoV-2 viral copies plus one, on a log-scale. Points represent the result for a single swab. Wi-fi plots show the peak daily number of simultaneous connections per building. No Wi-Fi data was available for the RES building. Shaded areas on the ‘copies’ and ‘wifi’ panels indicate university closures and study breaks.

We sought to evaluate surface detection of SARS-CoV-2 as a predictor of case burden at the building level. We applied a mixed-effects Poisson regression analysis; random intercepts were specified for buildings to account for repeated measurements. The incidence ratio, relating a 10-fold increase in the number of viral copies (plus one) to the total number of reported cases, was large and significant with a high level of certainty (IR = 17, 95% CI 7-44). IRs for individual sites ranged from 0.57 (ADM2) to 2.3 (FAC) with moderate variance among sites overall (SD = 0.62), but differences between sites were small relative to the uncertainty on their intercepts (Table S1).

### Surface detection of SARS-CoV-2 was not greater in high-traffic areas

At each building where environmental surveillance by surface swabbing was performed, we selected one high-traffic area where people often travel or congregate and a second low-traffic area for contrast. We hypothesized that the floors in commonly frequented locations would have greater rates of SARS-CoV-2 detection. However, positivity rates were similar across high-traffic (14.3%, N=280) and low-traffic sites (11.7%, N=274). To confirm this, we fit a mixed-effects logistic regression model, with the surface swab result as a binary outcome, the traffic level as a fixed effect, and specified random intercepts for each building to account for the clustering of sites. This model indicated that higher-traffic locations did not have significantly greater positivity rates than low-traffic locations (*OR = 1.3, 95% CI: 0.77 - 2.1*).

## DISCUSSION

In this study, we performed longitudinal environmental surveillance of SARS-CoV-2 across a university campus. We found that over the course of an academic year, detection of SARS-CoV-2 from built environment sampling sites across the campus correlated with reported campus-associated COVID-19 cases. Environmental detection of SARS-CoV-2 at university sites and regional SARS-CoV-2 wastewater signal were associated with an increase in campus-wide cases, according to a Poisson regression analysis. These results support other studies demonstrating that environmental sampling of SARS-CoV-2 RNA can be used to monitor COVID-19 cases at university campuses^9,10,21^.

The university setting provided a convenient test case for comparing the different scales of environmental testing. Built environment detection varied by building, with the residence hall associated with the highest burden (Fig. 3). Importantly, SARS-CoV-2 detection at two sites in January correlated with clusters of cases, indicating the usefulness of this method at identifying building-level case incidence. In contrast, a high built environment signal in the main library in January was not reflected in the case data, which could be an example of environmental surveillance identifying a location with unknown or unreported cases.

Despite evidence validating the use of environmental surveillance approaches, it is important to consider that factors specific to the study period may affect the accuracy of case trend projections. First, infection control measures were in place to reduce COVID-19 infections at the university including mandatory vaccination and masking for students, staff, and visitors to the University of Ottawa (Policy 129; https://www.uottawa.ca/about-us/policies-regulations), along with fewer campus in-person activities, smaller class sizes, and more on-line courses than in typical years. These mitigation measures likely contributed to the low COVID-19 case counts and low environmental detection observed in the Fall term. Second, the study period occurred during the global spread of the highly transmissible Omicron variant of SARS-CoV-2, which was likely responsible for the relatively large number of cases identified in January. The campus was closed for in-person learning in January 2022 to reduce spread of the Omicron variant. This would have limited the spatial distribution of cases on campus to buildings accessible to university members, such as the residence halls and administrative buildings.

For the above reasons, the specific trends in viral prevalence observed during the study period may be affected by pandemic-era mitigation measures and a unique episode of SARS-CoV-2 evolution. Future surveillance efforts can thus show what campus-wide trends occur after infection control measures are removed. In conclusion, our study validates the effectiveness of built environment surveillance to track SARS-CoV-2 prevalence in congregate settings, from university campuses to hospitals and long-term care homes^16,4^. At a practical level, environmental surveillance trends can be communicated to university community members to provide awareness of infectious disease risk on campus. Furthermore, as demonstrated for wastewater surveillance^22^, monitoring can be extended to other relevant infectious diseases (e.g., influenza), to provide a more complete picture of pathogen prevalence.

## Data Availability

All data produced in the present study are available upon reasonable request to the authors. Code and data will be made available from our GitHub repository upon publication.

## ACKNOWLEDGEMENTS

This study was funded by grants to RK from the University of Ottawa’s COVID-19 Recovery Task Force and the NSERC Alliance and Discovery Grants programs. We thank members of the Kassen lab, who generously volunteered to collect the swab samples over the course of the academic year.

## APPENDIX

**Table S1.** Summary of the random intercepts logistic regression model used for testing the effect of traffic-level (high vs. low traffic) on detection of SARS-CoV-2 in surface swabs by PCR.

| effect | group | term | estimate | std.error | statistic | p.value | conf.low | conf.high |
| --- | --- | --- | --- | --- | --- | --- | --- | --- |
| fixed |  | Intercept | 0.11 | 0.04 | -6.31 | < 0.0001 | 0.05 | 0.21 |
| fixed |  | traffic:high | 1.28 | 0.33 | 0.95 | 0.34 | 0.77 | 2.13 |
| ran_pars | site | SD (Intercept) | 0.71 |  |  |  |  |  |

